# Structural MRI Differences Between Parkinson’s Disease Motor Subtypes in Early-Stage: A Multicontrast Imaging Study

**DOI:** 10.1101/2024.12.08.24318615

**Authors:** Diógenes Diego de Carvalho Bispo, Edinaldo Gomes de Oliveira Neto, Pedro Renato de Paula Brandão, Danilo Assis Pereira, Talyta Cortez Grippe, Fernando Bisinoto Maluf, Neysa Aparecida Tinoco Regattieri, Andreia Faria, Xu Li, Maria Clotilde Henriques Tavares, Francisco Eduardo Costa Cardoso

## Abstract

**Background:** Parkinson’s disease (PD) is characterized by dopaminergic neuron degeneration, leading to motor and neuropsychological symptoms. PD is clinically divided into tremor-dominant (TD) and postural instability-gait disorder (PIGD) subtypes, which may differ in neuroanatomical changes. Neuroimaging explores these differences, enhancing understanding of PD heterogeneity.

**Objectives:** This study examines neuroanatomical differences between subtypes using MRI, focusing on subcortical volumes, cortical thickness, iron deposition, and white matter changes.

**Methods:** This cross-sectional study included 51 PD patients and controls. Participants underwent clinical assessments and MRI. Cortical and subcortical segmentation was automated using FreeSurfer, and quantitative susceptibility mapping was used to assess brain iron content. Diffusion-weighted MRI data were processed using Tractseg for tractometry analysis.

**Results:** The PD-TD group exhibited higher iron levels in the substantia nigra compared to healthy controls. Iron deposition in the thalamus correlated with MDS-UPDRS-part-III and PIGD scores. Tractometry showed differences in fractional anisotropy (FA) between PD-TD and PD-PIGD in the bilateral fronto-pontine tract (FPT). The PD-PIGD group had decreased FA in the middle cerebellar peduncle (MCP) compared to controls. FA in the left FPT correlated with tremor scores, while FA in the MCP correlated with PIGD scores.

**Conclusions:** This study highlights distinct neuroimaging signatures between PD motor subtypes. Elevated iron deposition in the substantia nigra is a shared feature, particularly in the TD subtype. Subtype-specific white matter changes, including reduced FA in the FPT and MCP, correlate with tremor and PIGD scores. These findings underscore the potential of neuroimaging biomarkers in unraveling PD heterogeneity and guiding tailored approaches.

## INTRODUCTION

Parkinson’s disease (PD) is a complex neurodegenerative disorder characterized by a constellation of motor and non-motor symptoms.[1] The cardinal motor features of PD include bradykinesia, rigidity, resting tremor, and postural instability, which are primarily attributed to the loss of dopaminergic neurons in the substantia nigra pars compacta and the consequent depletion of striatal dopamine.[2] However, the clinical presentation of PD is remarkably heterogeneous, leading to the recognition of distinct motor subtypes, namely tremor-dominant (TD) and postural instability-gait disorder (PIGD).[3]

The TD subtype is characterized by the predominance of tremor symptoms, while the PIGD subtype is dominated by axial symptoms such as postural instability, gait difficulties, and freezing of gait.[4] These subtypes have been associated with different clinical courses, cognitive profiles, and responses to treatment, suggesting that they may have distinct underlying pathophysiological mechanisms.[5,6]

Neuroimaging studies have provided valuable insights into the structural and functional brain alterations associated with PD motor subtypes. Magnetic resonance imaging (MRI) has revealed differences in cortical thickness, subcortical volumes, and white matter integrity between TD and PIGD subtypes.[7,8] Furthermore, advanced MRI techniques such as quantitative susceptibility mapping (QSM) have demonstrated differential patterns of iron deposition in the basal ganglia, cerebellar dentate and substantia nigra in PD subtypes.[9] Despite these findings, the neuroanatomical correlates of PD motor subtypes remain incompletely understood,[10] and there is a need for comprehensive, multicontrast MRI studies to better characterize the distinct pathophysiological signatures of these subtypes. Such studies could potentially identify imaging biomarkers that could aid in the differential diagnosis, prognosis, and personalized treatment of PD subtypes.

In this context, the present study aimed to investigate the neuroanatomical differences between PD motor subtypes using a multicontrast MRI approach. Specifically, we employed QSM to assess iron deposition, diffusion tensor imaging (DTI) and tractometry to evaluate white matter integrity, and T1-weighted imaging to examine cortical thickness and subcortical volumes. We hypothesized that TD and PIGD subtypes would exhibit distinct patterns of iron deposition, white matter alterations, and structural brain changes, which would correlate with their clinical characteristics. By providing a comprehensive neuroimaging characterization of PD motor subtypes, this study seeks to advance our understanding of the pathophysiological underpinnings of PD heterogeneity and potentially guide the development of targeted therapeutic strategies.

## METHODS

### Ethical compliance statement

This study was conducted in accordance with the ethical principles outlined in the Declaration of Helsinki and its subsequent revisions. The study protocol was approved by the Research Ethics Committee of the Centro Universitário de Brasília (CEUB) (Approval No. 07073419.0.0000.0023). All participants provided written informed consent prior to their inclusion in the study. The consent process involved a detailed explanation of the study objectives, procedures, potential risks and benefits, and the voluntary nature of participation. Participants were given adequate time to review the consent form and ask questions before making their decision. The study team ensured that all participants had the capacity to provide informed consent and that their privacy and confidentiality were protected throughout the study.

### Study design and context

This was a prospective, analytical, cross-sectional study conducted at a single center, the Neuroscience and Behavior Laboratory at the University of Brasília, Campus Darcy Ribeiro. Recruitment commenced in the first half of 2019 and concluded in August 2021. Due to the COVID-19 pandemic, enrollment was temporarily suspended for most of 2020 to minimize the risk of SARS-CoV-2 transmission in the research setting.

### Participants

Patients with PD were prospectively recruited from the community through a combination of referrals from attending physicians and active outreach by the research team following public disclosure of the study on social media and television interviews. This recruitment strategy aimed to enhance the generalizability of the study findings.

The inclusion criteria for PD patients were based on the Queen Square Brain Bank (QSBB) diagnostic criteria,[11] with the exception of the “family history” item. Patients were excluded if they met any of the following criteria: (a) clinically defined dementia, according to consensus standards;[12] (b) fulfillment of any of the Movement Disorder Society (MDS) exclusion criteria for PD diagnosis;[13] (c) inability to understand Portuguese; (d) advanced chronic organic disease, as assessed by the principal investigator; (e) claustrophobia; (f) pregnancy; or (g) presence of a cardiac pacemaker, intracranial aneurysm clip, or deep brain stimulation electrode, which are contraindications for MRI.

### Clinical assessment

All patients underwent a comprehensive clinical evaluation during the “on” levodopa state. Motor signs were assessed by a single neurologist (PB) using the Movement Disorder Society Unified Parkinson’s Disease Rating Scale (MDS-UPDRS) [14] and the Hoehn & Yahr (H&Y) staging system.[15] Demographic and clinical data, including years of formal education, age at disease onset, current medications, and dosages, were collected during patient interviews. All participants were on stable dopaminergic medication regimens for at least one month prior to study enrollment. Global cognitive function was evaluated using the Parkinson’s Disease-Cognitive Rating Scale (PD-CRS)[16,17] and the Montreal Cognitive Assessment (MoCA).[18,19]

Parkinson’s disease motor subtypes were classified based on the ratio of mean MDS-UPDRS tremor scores to mean MDS-UPDRS postural instability and gait difficulty scores, yielding three categories: (a) TD, (b) PIGD, and (c) intermediate subtype. The MDS-UPDRS-derived cut-off scores for subtype classification were as follows: TD > 1.15 and PIGD < 0.90.[20] The tremor score was calculated by summing 11 items from the MDS-UPDRS (items 2.10 and 3.15–3.18), while the PIGD score was derived from the sum of five items (items 2.12: walking and balance, 2.13: freezing, 3.10: gait, 3.11: freezing of gait, and 3.12: postural stability).

### MRI data acquisition

MRI was performed using a Philips Achieva 3T scanner (Best, Netherlands) equipped with an 8-channel SENSE receive coil. The following MRI sequences were obtained: (a) Three dimensional (3D) T1-weighted sequence, turbo field echo, sagittal, with field of view (FOV) = 208 × 240 × 256 mm^3^, reconstructed resolution of 1 × 1 × 1 mm^3^, echo time (TE) = min full echo, repetition time (TR) = 2,300 ms, TI = 900 ms, two times accelerated acquisition (the acquisition is accelerated by a factor of two, taking a total time of 4 minutes and 52 seconds); (b) Three-dimensional gradient-recalled echo (GRE) multiecho sequence with a resolution of 0.5 × 0.5 × 2.0 mm^3^, transverse orientation, TE1/dTE = 6.0 ms/7.3 ms, TR = 50 ms, number of echoes = 5, FOV = 240 × 240 mm, flip angle = 20 degrees, bandwidth = 150 Hz/pixel (the acquisition was accelerated 1.8 times, with a total scan time of 7 minutes and 52 seconds); and (c) Diffusion-weighted sequence, axial, with FOV 232 × 232 × 160 mm^3^, reconstructed resolution of 2 × 2 × 2 mm^3^, TE = 71 ms; TR = 3,300 ms, 32 diffusion encoding directions (b = 800 s/mm^2^), (total acquisition time of 6 minutes and 46 seconds). The 3D GRE multiecho sequence was adapted from the Magnetic Resonance Research Laboratory at Cornell University [https://pre.weill.cornell.edu/mri/pages/qsm.html]. The remaining sequences were adjusted according to the Alzheimer’s Disease Neuroimaging Initiative (ADNI) protocol [https://adni.loni.usc.edu/data-samples/adni-data/neuroimaging/mri/mri-scanner-protocols/].

### MRI data processing

The processing of MRI was carried out in three stages as shown in Figure 1.

**Figure 1.**
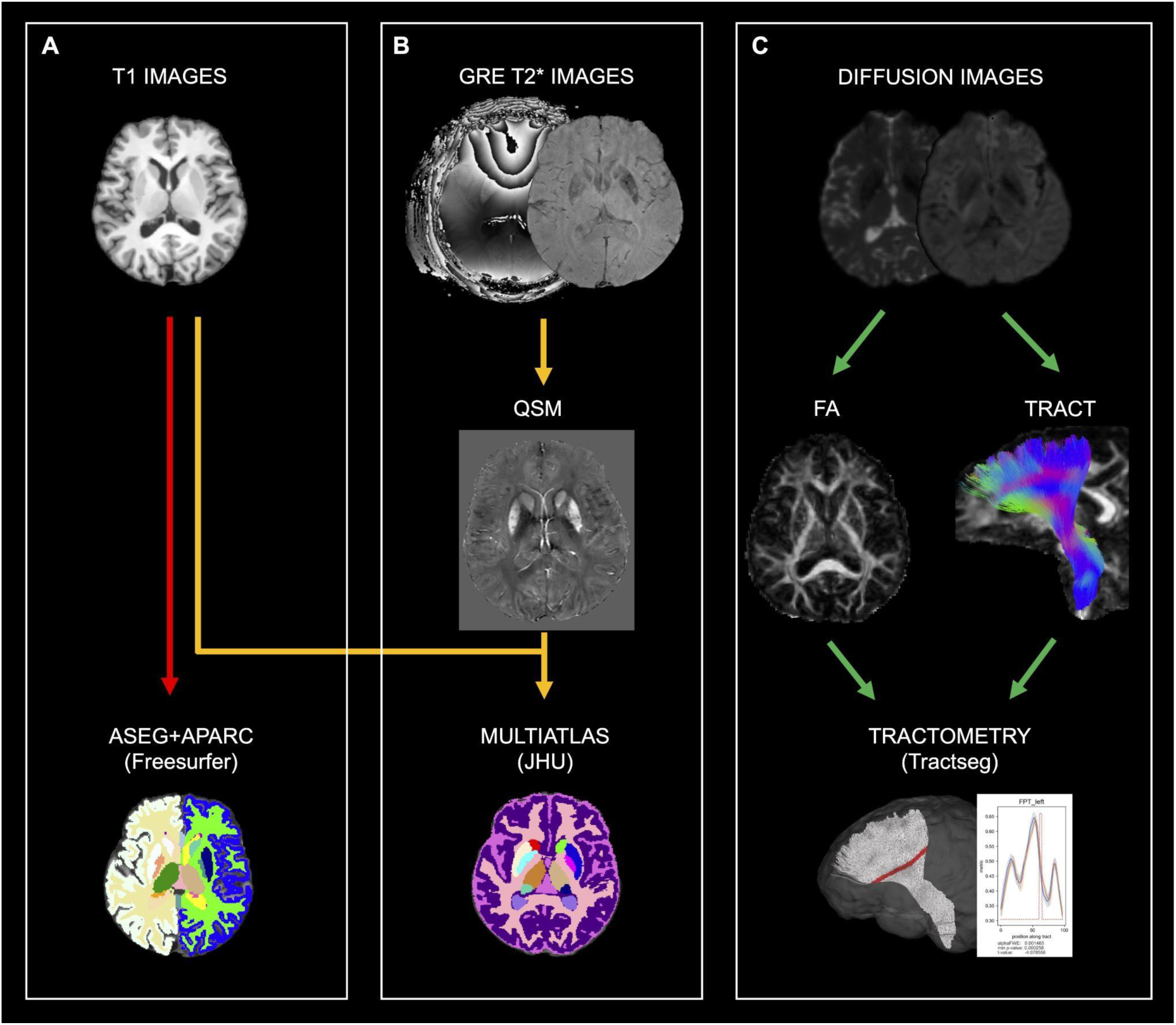
MRI processing pipelines. (A) Raw T1-weighted images undergo gray matter segmentation using the FreeSurfer suite. (B) QSM was derived from both magnitude and phase images acquired with the T2*-weighted GRE sequence and, together with T1-weighted images, was analyzed for brain iron and subcortical structural volumes using the QSM Toolbox and JHU multi-atlas matching. (C) Diffusion MRI-derived measures including Fractional Anisotropy (FA) are computed, and bundle-specific tractography is performed using Tractseg. Tractometry of each bundle using the FA is conducted.

#### Automated cortical and subcortical segmentation

Cortical thickness and deep gray matter (GM) nuclei volume were estimated using the FreeSurfer software suite (version 7.1)[21] on T1-weighted images. The analysis pipeline included several steps: (1) bias correction to account for spatial variations in image intensity; (2) Normalization of the brain to a standard Talairach coordinate system; (3) segmentation of white matter (WM) and deep GM structures; (4) triangular tessellation of the WM-GM boundary; and (5) surface warping by tracking intensity gradients to accurately identify the WM-GM and GM-pial borders. The cortical surface was then inflated, followed by spherical atlas registration and parcellation into gyral and sulcal structures based on the Desikan-Killiany atlas. Cortical thickness was calculated as the distance between the GM-WM boundary and the corresponding pial surface at each vertex. A 10 mm full width at half maximum (FWHM) symmetric Gaussian kernel was applied to smooth the cortical thickness data, which helps to reduce noise and improve signal-to-noise ratio.

Subcortical and limbic structure volumes were automatically segmented by assigning a neuroanatomical label to each voxel in the MRI volume based on probabilistic information derived from a manually labeled training set. The following structures were segmented bilaterally: caudate, putamen, globus pallidus, hippocampus, nucleus accumbens, and amygdala. To account for individual differences in head size, the volumes were normalized to the estimated total intracranial volume. For statistical analysis, the volumes of the right and left hemispheres were summed to obtain a single value for each structure.

#### Quantitative Susceptibility Mapping

QSM was used to assess brain iron content. QSM images were reconstructed using the Johns Hopkins University/Kennedy Krieger Institute QSM Toolbox, version 3.0 [http://godzilla.kennedykrieger.org/QSM/]. The processing pipeline included several steps. First, the MR phase images acquired with the 3D GRE sequence were unwrapped using a best-path-based method.[22] This unwrapped phase image was then combined with the FSL brain extraction tool applied to the third-echo GRE magnitude image with one voxel erosion to create a brain mask.[23] Background field was removed using the Variable-kernel Sophisticated Harmonic Artifact Reduction for Phase (V-SHARP) method with a maximum kernel size of 6 mm and a regularization parameter of 0.05.[24,25] The resulting tissue field images were averaged across all echoes to achieve a higher SNR compared to single-echo reconstruction.[26]

Quantitative susceptibility maps were generated using the modified Structural Feature-based Collaborative Reconstruction method with automatic referencing (SFCR+0), which is a modified version of the QSM approach.[27] This method exploits the structural consistency between the magnitude and susceptibility images to improve the reconstruction quality. The QSM values were referenced to automatically selected central cerebrospinal fluid (CSF) regions based on a small effective transverse relaxation rate (R2* < 5 Hz) to exclude choroid plexus regions which may contain iron or calcium deposition.[28]

#### QSM Image Segmentation and Quantification

The procedures for obtaining regions of interest (ROIs) based quantification of tissue magnetic susceptibility in the deep gray matter were summarized as follows. First, the T1-weighted image was coregistered to the GRE magnitude image acquired at TE of 6 ms using FSL FLIRT.[29] The coregistered T1 and QSM images were then segmented using a multicontrast multiatlas matching approach, which is developed as part of the Johns Hopkins University brain atlas [mricloud.org].[30,31] Twenty ROIs (caudate, putamen, internal and external globus pallidus, thalamus, pulvinar, substantia nigra, subthalamic nucleus, red nucleus, and dentate) were automatically segmented for quantifying tissue magnetic susceptibility (iron measure) in each region. In comparative analyses, the iron measures of the right and left structures were summed.

#### Diffusion-weighted MRI processing

Diffusion-weighted MRI (dMRI) data were processed using TractoFlow,[32] an automated pipeline that includes preprocessing steps such as motion correction, geometric distortion correction, and bias field correction. The preprocessed dMRI data were then fitted with a diffusion tensor model using FSL-DTIFIT to estimate the three eigenvalues and eigenvectors at each voxel. Fractional anisotropy (FA) maps were calculated from the eigenvalues to quantify the degree of anisotropic water diffusion in the brain.

Along-tract FA statistics (tractometry) were generated using TractSeg,[33,34] a machine learning-based tool that creates tract masks and orientation maps for accurate bundle-specific tractography. The tractometry analysis followed the methodology described by Chandio.[35] First, each streamline was resampled to 100 equidistant points, and the FA values were calculated at each of these points. Next, the QuickBundles algorithm[36] was used to identify the centroid streamline for each tract. Each resampled streamline point was then assigned to the nearest centroid point. Finally, the FA values were averaged across all streamline points assigned to each centroid point, resulting in 100 FA measurements along the length of each tract.

To reduce the risk of false-positive findings, the tractometry analysis focused on a subset of motor projection and cerebellar tracts: corticospinal tract (CST), fronto-pontine tract (FPT), inferior cerebellar peduncle (ICP), middle cerebellar peduncle (MCP), superior cerebellar peduncle (SCP), thalamo-premotor tract (T-PREM), and striato-premotor tract (ST-PREM). These tracts were selected based on their relevance to motor function and potential involvement in Parkinson’s disease.[37,38]

The use of automated processing tools like TractoFlow and TractSeg ensures the reproducibility and efficiency of the dMRI analysis. The preprocessing steps help mitigate common artifacts and improve data quality. The diffusion tensor model provides a simple yet robust characterization of water diffusion in the brain, with FA serving as a sensitive measure of microstructural integrity. The tractometry approach, which combines machine learning-based segmentation with along-tract analysis, enables the detection of localized FA changes within specific white matter pathways. By focusing on a limited number of tracts, the analysis strikes a balance between comprehensive examination and statistical power, reducing the likelihood of spurious findings. These advanced dMRI processing techniques contribute to a more reliable and meaningful investigation of white matter alterations in Parkinson’s disease subtypes.

#### MRI quality control

All raw and processed MRI datasets were carefully inspected for artifacts, including gross geometric distortion, bulk motion, and signal dropout. Quality control for T1-weighted and diffusion-weighted MRI (dMRI) data was performed using Dmriqc-flow, a comprehensive quality control tool for dMRI.[39] Additionally, a board-certified neuroradiologist visually reviewed the cortical and subcortical segmentations and white matter tracts to ensure their accuracy and adherence to anatomical boundaries. Datasets with significant artifacts or segmentation errors were excluded from further analysis to maintain the integrity and reliability of the results.

### Statistical analysis

#### Demographic, clinical, and cognitive assessments

Statistical analyses were conducted using R software (version 4.4.0, R Foundation for Statistical Computing, Vienna, Austria, 2024) and its associated packages. Depending on the nature of the variables and the assumptions of the tests, group comparisons for clinical and demographic characteristics were performed using Student’s t-tests, Mann-Whitney U-tests, chi-square tests, or analysis of covariance (ANCOVA) as appropriate. These tests allowed for the identification of potential differences between Parkinson’s disease subtypes and healthy controls while controlling for relevant confounding factors.

#### Analysis of cortical thickness and subcortical volume

Vertex-wise cortical thickness was analyzed using generalized linear models (GLM) for each hemisphere separately. Group comparisons between Parkinson’s disease patients and healthy controls were conducted using the “mri_glmfit” function in FreeSurfer.[21] Multiple comparisons correction was performed using Monte Carlo simulations with a cluster-forming threshold of p < 0.001. Age was included as nuisance covariate to account for their potential influence on cortical thickness. For the analysis of subcortical gray matter nuclei volumes, ANCOVA was employed to compare the groups while controlling for age, sex, and estimated total intracranial volume. Post hoc Bonferroni pairwise tests were used to correct for multiple comparisons and identify specific group differences.

#### ROI-Based Subcortical QSM Analysis

Subcortical gray matter nuclei QSM values were subjected to ANCOVA, with age included as a covariate to control for its potential confounding effects. To rigorously address multiple comparisons and isolate significant intergroup differences, post hoc pairwise comparisons were performed using Bonferroni correction.

#### Analysis of Tractometry

Correlations and T-tests were applied point-wise on the along-tract FA. We used the permutation-based multiple comparison correction (with n = 5000 repetitions) published by Nichols and Holmes (2001)[40,41] to appropriately adjust p values given the correlation structure of the data. We used an uncorrected confidence threshold of p < 0.05 (the corrected p value is different for each bundle depending on its correlation structure). Controlling for age and sex covariates was done by regressing them out of the data before calculating the t-test (categorical approach) or correlation (dimensional approach).

## RESULTS

### Demographic and Clinical Characteristics

A total of 77 participants were initially recruited for the study. In the Parkinson’s disease (PD) group, 26 participants were excluded for the following reasons: dementia (n=3), history of stroke (n=3), progressive supranuclear palsy (n=2), lack of MRI exams (n=17), and low-quality MRI exam (n=1) (Figure 2).

**Figure 2.**
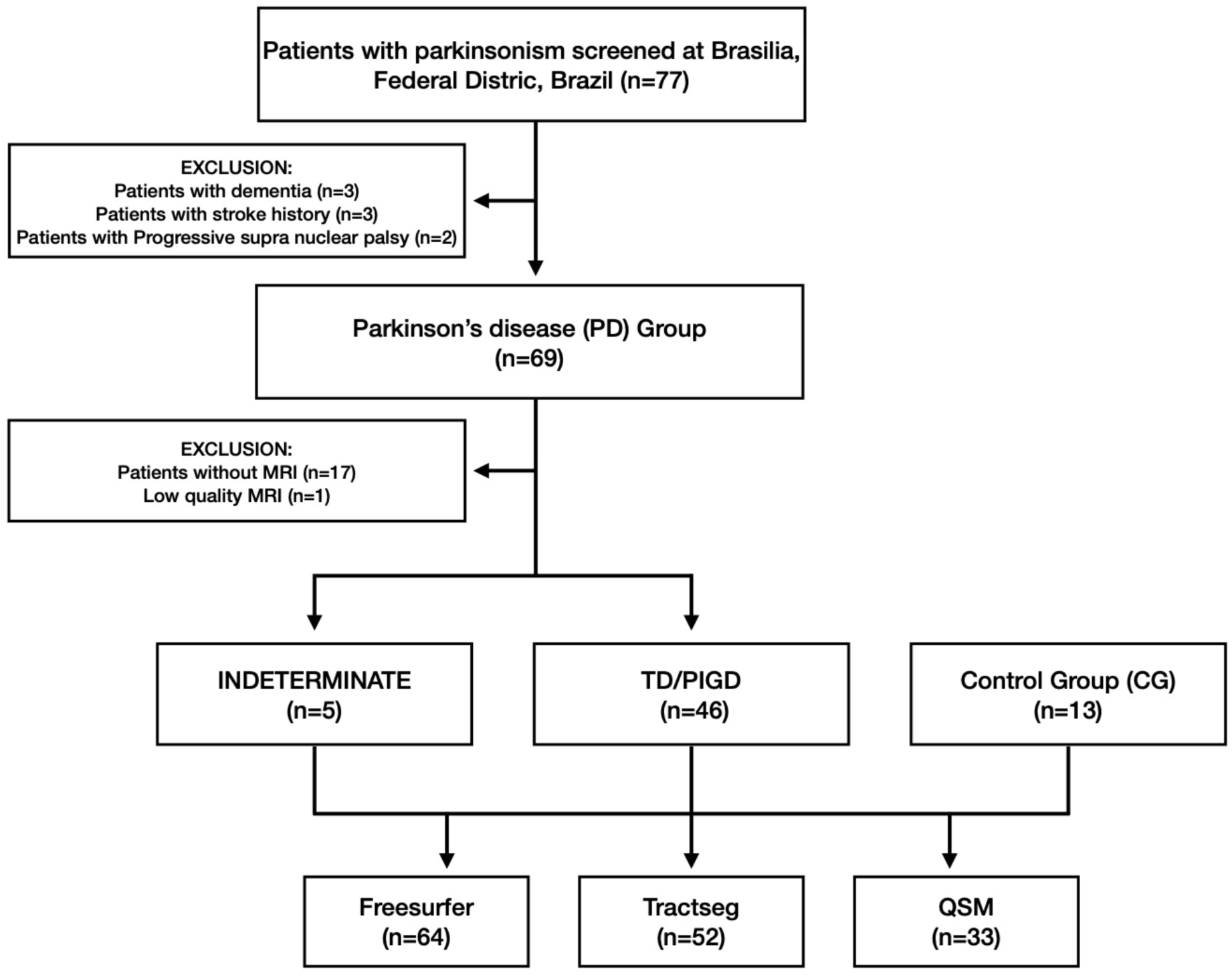
Flowchart with the enrollment of participants in the PD and Control groups and the investigations that were carried out.

**Figure 3.**
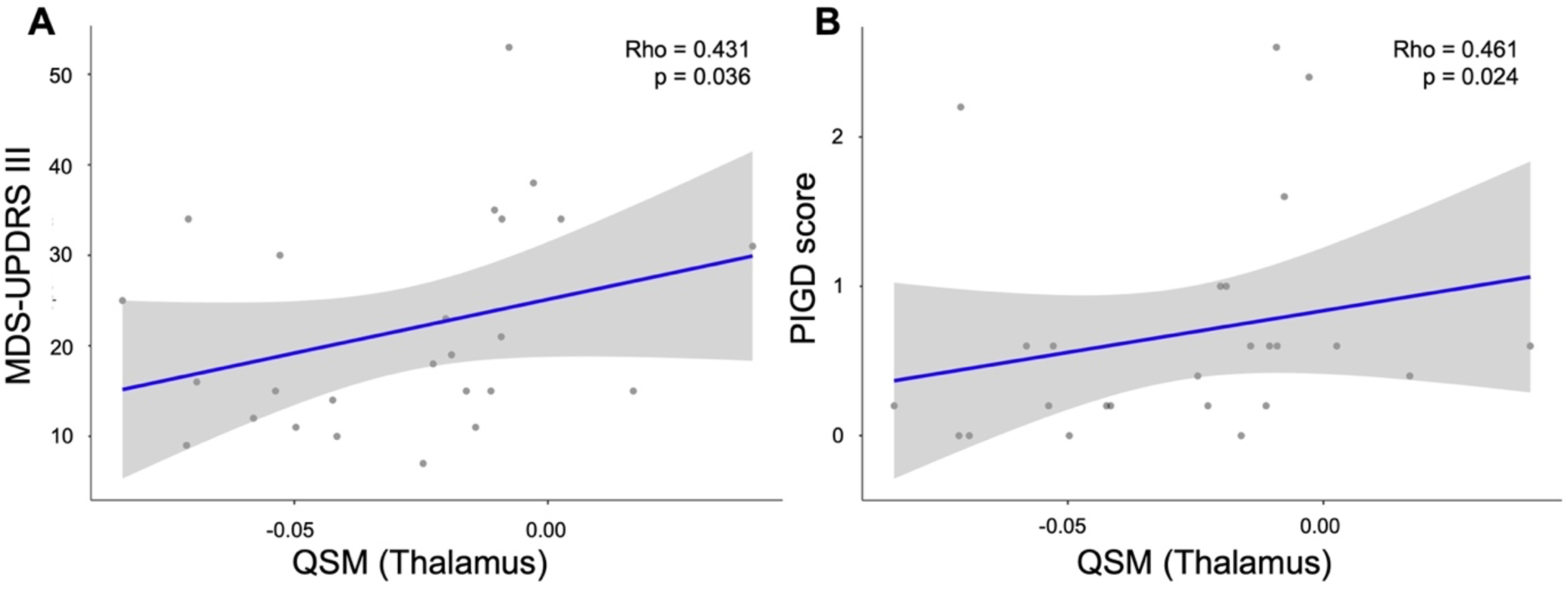
Correlation analysis between evaluation scales and susceptibility values in the PD group with age as covariable. (A) Partial correlation between MDS-UPDRS III score and thalamus QSM. (B) Partial correlation between PIGD score and thalamus QSM. N = 25.

The remaining 51 PD patients were classified into three groups based on their motor subtypes: TD (n=23, 45.1%), PIGD (n=23, 45.1%), and an intermediate group (n=5, 9.8%). Due to its small sample size and lack of predominant motor symptoms, the intermediate group was excluded from intergroup comparisons but included in the correlation analyses. Additionally, 13 age- and sex-matched healthy controls (HCs) were included in the study. No significant differences were observed among the groups in terms of sex, age, or years of education. The MoCA and PD-CRS scores also did not differ significantly between the PD subgroups and the control group. However, the TD group had significantly lower H&Y stages and MDS-UPDRS part III scores compared to the PIGD group (Table 1).

**Table 1.** Demographic and clinical features (TD, PIGD, and Control groups).

|  | TD | PIGD | Control |  |
| --- | --- | --- | --- | --- |
| Characteristics | (n=23, 39%) | (n= 23, 39%) | (n= 13, 22%) | Statistic |
| Age (y) | 61.8 ± 11.5 (37, 76) | 58.9 ± 10.4 (44, 80) | 63.6 ± 8.0 (51, 77) | F = 0.95; p = 0.391<sup>a</sup> |
| Sex |  |  |  | X<sup>2</sup> = 1.59; p = 0.453<sup>b</sup> |
| Male, n (%) | 16 (69.6%) | 17 (73.9%) | 7 (53.9%) |  |
| Female, n (%) | 7 (30.4%) | 6 (26.1%) | 6 (46.1%) |  |
| Education (y) | 16.0 ± 4.7 (8, 30) | 14.6 ± 5.0 (3, 22) | 17.0 ± 3.1 (11, 21) | F = 1.28; p = 0.285<sup>a</sup> |
| MoCA | 25.7 ± 2.5 (20, 29) | 24.0 ± 3.6 (16, 30) | 26.4 ± 2.6 (21, 29) | F = 2.36; p = 0.104<sup>c</sup> |
| PDCRS | 98.0 ± 13.4 (66, 117) | 93.6 ± 16.4 (69, 120) | 101 ± 10.5 (78, 111) | F = 1.70; p = 0.192<sup>c</sup> |
| Disease duration (y) | 3.3 ± 3.0 (1,14) | 4.7 ± 3.1 (1, 13) |  | U = 351; p = 0.056<sup>d</sup> |
| Hoehn yahr | 1.3 ± 0.5 (1, 2) | 1.74 ± 0.5 (1, 3) |  | U = 158; p = 0.008<sup>d</sup> |
| MDS-UPDRS III | 19.7 ± 9.6 (8, 45) | 26.7 ± 12.8 (7, 53) |  | t = -2.11; p = 0.041<sup>e</sup> |
| Tremor score | 0.7 ± 0.4 (0.1, 1.7) | 0.3 ± 0.2 (0, 0.7) |  | U = 76.5; p = <0.001<sup>d</sup> |
| PIGD score | 0.2 ± 0.2 (0, 0.6) | 1.0 ± 0.7 (0.2, 2.6) |  | U = 27.5; p = <0.001<sup>d</sup> |

Abbreviations: MoCA, Montreal Cognitive Assessment;
Data are shown as mean ± standard deviation (minimum, maximum) or n (%).
<sup>a</sup>ANOVA one way: TD, PIGD, and Control
<sup>b</sup>X<sup>2</sup> test: TD, PIGD, and Control
<sup>c</sup>ANCOVA: TD, PIGD, and Control, correcting for age and education.
<sup>d</sup>Mann-Whitney test: TD and PIGD
<sup>e</sup>Independent t-test: TD and PIGD

**Table 2.** Comparison of the volumes of subcortical structures between TD, PIGD, and Control group.

|  | TD | PIGD | Control |  |  |
| --- | --- | --- | --- | --- | --- |
| Regions | (n=23, 39%) | (n= 23, 39%) | (n= 13, 22%) | p value<sup>a</sup> | Post hoc<sup>b</sup> |
| Thalamus | 0.953 ± 0.0169 | 0.958 ± 0.0171 | 1.022 ± 0.0227 | p = 0.039 | TD vs Control p = 0.050 |
| Caudate | 0.426 ± 0.0107 | 0.436 ± 0.0108 | 0.453 ± 0.0143 | p = 0.316 |  |
| Putamen | 0.589 ± 0.0115 | 0.623 ± 0.0116 | 0.621 ± 0.0154 | p = 0.087 |  |
| Pallidum | 0.258 ± 0.0040 | 0.264 ± 0.0040 | 0.258 ± 0.0054 | p = 0.598 |  |
| Hippocampus | 0.521 ± 0.0112 | 0.518 ± 0.0113 | 0.560 ± 0.0151 | p = 0.068 |  |
| Amygdala | 0.221 ± 0.0056 | 0.225 ± 0.0056 | 0.235 ± 0.0075 | p = 0.322 |  |
| Accumbens area | 0.063 ± 0.0022 | 0.066 ± 0.0022 | 0.069 ± 0.0029 | p = 0.152 |  |

Data are shown as mean ± standard deviation, in the unit of mm<sup>3</sup>
<sup>a</sup>ANCOVA: TD, PIGD, and Control, correcting for age
<sup>b</sup>Post hoc Bonferroni pairwise test

**Table 3.** Comparison of the susceptibility values (QSM) between TD, PIGD, and Control group.

|  | TD | PIGD | Control |  |  |
| --- | --- | --- | --- | --- | --- |
| Regions | (n=10, 33%) | (n= 12, 40%) | (n= 8, 27%) | p value<sup>a</sup> | Post hoc<sup>b</sup> |
| **Caudate** | 0.117 ± 0.0098 | 0.126 ± 0.0091 | 0.120 ± 0.0113 | p = 0.766 |  |
| **Dentate** | 0.180 ± 0.0186 | 0.196 ± 0.0172 | 0.171 ± 0.0214 | p = 0.660 |  |
| **Globus pallidus external** | 0.356 ± 0.0158 | 0.330 ± 0.0147 | 0.322 ± 0.0182 | p = 0.329 |  |
| **Globus pallidus internal** | 0.277 ± 0.0156 | 0.266 ± 0.0145 | 0.232 ± 0.0180 | p = 0.170 |  |
| **Putamen** | 0.156 ± 0.0130 | 0.155 ± 0.0120 | 0.170 ± 0.0149 | p = 0.733 |  |
| **Red nucleus** | 0.192 ± 0.0176 | 0.196 ± 0.0163 | 0.171 ± 0.0202 | p = 0.614 |  |
| **Substantia nigra** | 0.300 ± 0.0220 | 0.257 ± 0.0204 | 0.195 ± 0.0253 | p = 0.015 | TD vs Control p = 0.012 |
| **Subthalamic nucleus** | 0.195 ± 0.0178 | 0.176 ± 0.0165 | 0.155 ± 0.0204 | p = 0.359 |  |
| **Thalamus** | -0.042 ± 0.0101 | -0.025 ± 0.0094 | -0.038 ± 0.0116 | p = 0.458 |  |

Data are shown as mean ± standard deviation, in the unit of ppm
<sup>a</sup>ANCOVA: TD, PIGD, and Control, correcting for age
<sup>b</sup>Post hoc Bonferroni pairwise test

### Cortical thickness and subcortical structures volume

The vertex-wise analysis of cortical thickness revealed no significant differences between the groups. Although a significant difference in thalamic volume was found when comparing the TD, PIGD, and HC groups, post hoc analyses did not yield any significant pairwise differences.

### Susceptibility Values

Significant differences in QSM values were found in the substantia nigra between the PD and control groups. Post hoc analysis revealed a significant difference between the TD and control groups (p = 0.012).

In the PD group, partial correlations were observed between QSM values and clinical scores, controlling for age as a covariate. The QSM values in the thalamus showed significant partial correlations with the MDS-UPDRS part III score (Rho = 0.431, p = 0.036) and the PIGD score (Rho = 0.461, p = 0.024).

### Tractometry

Using along-tract statistics derived from TractSeg, no significant differences in FA were found between the TD group and the control group. However, compared to the control group, the PIGD group exhibited significantly decreased FA in the MCP, right SCP, right ICP, and right ST-PREM (Figure 4). Significant differences in FA were also observed between the TD and PIGD groups in the bilateral FPT (with TD showing lower FA). Additionally, the PIGD group showed significantly decreased FA in the MCP compared to the TD group (Figure 5).

**Figure 4.**
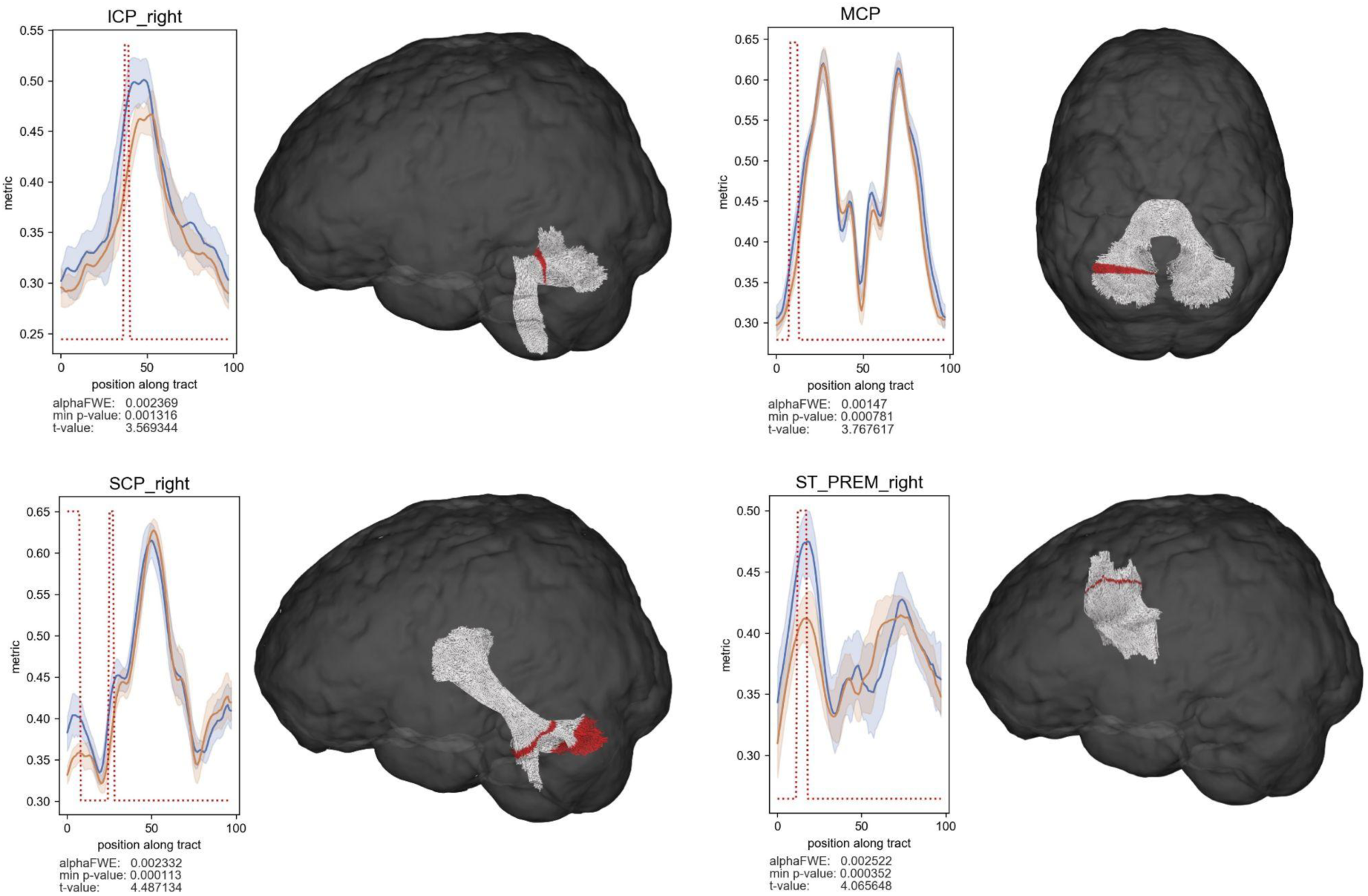
Tractometry results for the group analysis of control and PIGD patients. The blue (control) and orange (PIGD) line show the mean FA along each tract for the respective group and the 95% confidence interval. The red dotted line indicates in a binary way at which positions along the tract there are differences between the two groups. N=36.

**Figure 5.**
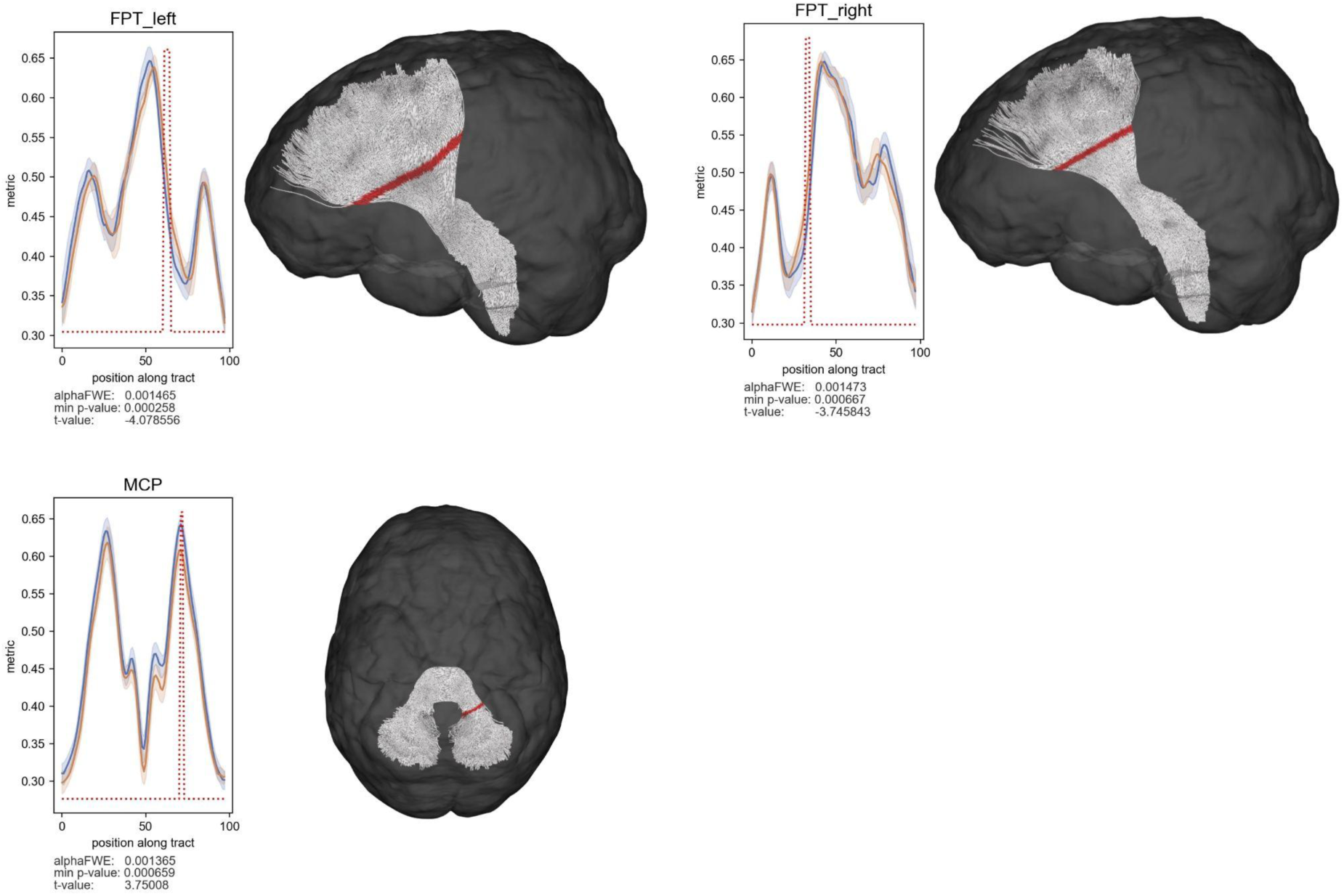
Tractometry results for the group analysis of TD and PIGD patients. The blue (TD) and orange (PIGD) line show the mean FA along each tract for the respective group and the 95% confidence interval. The red dotted line indicates in a binary way at which positions along the tract there are differences between the two groups. N=36.

In the PD group, significant correlations were found between the tremor score and FA in the left FPT, and between the PIGD score and FA in the MCP (Figure 6). Detailed tractometry results for all analyzed tracts are provided in the supplementary material.

**Figure 6.**
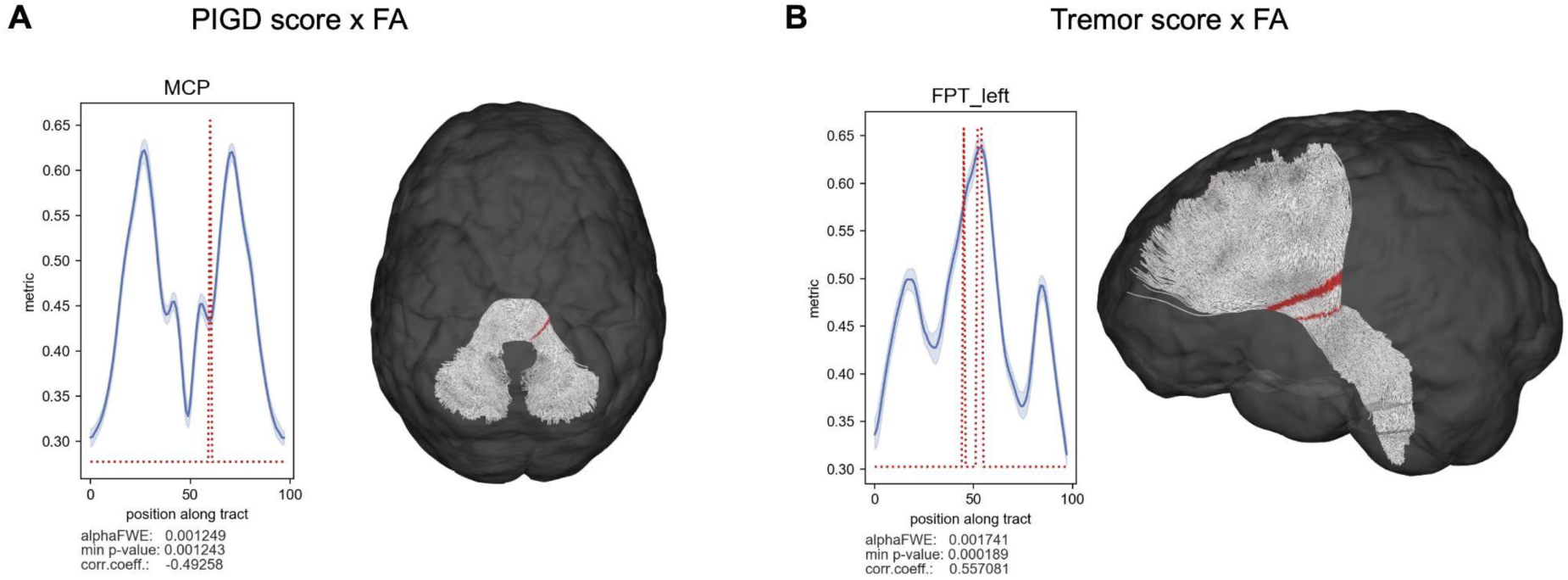
Tractometry results for the correlation analysis between PIGD score or tremor score and FA in the PD group with age and sex as covariable. The blue line shows the mean FA along each tract for the PD group and the 95% confidence interval. The red dotted line indicates in a binary way at which positions along the tract there are significant correlations. N = 40.

## DISCUSSION

Our multicontrast MRI study identified distinct neuroimaging signatures between different motor subtypes of PD, offering important insights into the neuropathological basis of PD heterogeneity. While there were no significant differences in subcortical volumes or cortical thickness between PD subtypes and healthy controls, increased iron deposition in the substantia nigra was observed in PD patients, particularly in those with the TD subtype. Tractometry analysis revealed more pronounced alterations in white matter integrity in the PIGD subtype compared to both controls and the TD subtype, especially in cerebellar pathways. These findings suggest that cerebellar circuits play a crucial role in postural and gait dysfunctions, contributing to the distinct motor phenotypes observed in PD.

The elevated iron content in the substantia nigra of PD patients, as measured by QSM, aligns with the well-established pathophysiology of PD, characterized by the progressive loss of dopaminergic neurons and subsequent iron accumulation.[42] In our study, we observed increased iron deposition in the substantia nigra of the TD group compared to the control group, suggesting a potential role of iron accumulation in the development of tremor symptoms in early-stage Parkinson’s disease. This finding is consistent with previous studies,[43,44][44] which highlights iron deposition in the substantia nigra as a possible biomarker for PD. Notably, studies have shown that iron deposition initially accumulates in the substantia nigra pars compacta (SNpc) during the early stages of the disease, with subsequent progressive dispersion to other neural regions. Furthermore, a correlation has been identified between iron accumulation in the dentate nucleus and the severity of tremor symptoms.[45,46] However, it is important to note that the QSM atlas used in our study did not differentiate between the SNpc and substantia nigra pars reticulata (SNpr), which may account for some discrepancies in our results.

The tractometry findings offer valuable insights into the distinct white matter alterations among PD motor subtypes. While no significant differences in FA were detected between the TD group and healthy controls, the PIGD group showed marked reductions in FA across several cerebellar tracts. This suggests that the PIGD subtype is characterized by more extensive disruptions in white matter integrity, particularly in cerebellar pathways involved in motor coordination. The significant correlation between PIGD scores and FA in the MCP further supports the cerebellum’s central role in PD motor subtypes. A study highlights brain network alterations in PIGD patients, revealing less efficient communication between regions such as the cerebellar vermis and prefrontal cortex. These changes are associated with more severe motor symptoms, indicating that structural brain differences may underlie the pronounced motor decline in PIGD patients.[47] Additionally, another study found cerebellar atrophy and altered brain activity in PIGD patients during walking-simulation tasks. These patients exhibited increased activation in cognitive regions of the cerebellum, likely as compensation for diminished motor function.[48] Such cerebellar alterations may contribute to their motor and gait impairments. The evolving involvement of the cerebellum at various disease stages underscores the complexity and dynamic nature of PD pathophysiology.[49–52]

The TD group exhibited lower FA in the bilateral FPT compared to the PIGD group, suggesting that tremor-dominant PD may involve disruptions in the cortico-cerebellar circuit, contributing to tremor pathophysiology. The significant correlation between tremor severity and FA in the left FPT further underscores that specific tract alterations are closely linked to the intensity of motor symptoms in PD. These findings highlight the potential of tractometry to identify structural correlates of motor subtype-specific impairments in Parkinson’s disease. Emerging evidence also highlights the importance of the cortico-thalamo-cerebellar circuit in motor function, both in healthy individuals and those with motor dysfunctions. The interaction between the cerebellum and other motor pathways may play a key role in shaping the clinical presentation of PD, particularly the tremor commonly observed in the TD subtype.[51,53,54] Moreover, the FPT is a crucial white matter pathway that connects the frontal lobe to the pons. It facilitates communication between the frontal cortex, which is responsible for higher-order motor planning, and the cerebellum, which fine-tunes and coordinates movements. In the TD subtype of PD, alterations in the FPT likely reflect impaired communication between these regions, contributing to the characteristic tremor. This disruption of the cortico-cerebellar circuit may interfere with the regulation and execution of precise motor actions, further exacerbating tremor symptoms in TD patients.[55–57]

While our study provides valuable insights into the neuroimaging correlates of PD motor subtypes, several limitations should be acknowledged. There are challenges associated with QSM that warrant attention. QSM measures the overall magnetic susceptibility of tissue, which can reflect contributions from both iron and myelin, especially in regions like the thalamus. This lack of specificity could impact the interpretation of our findings. However, recent advancements in QSM, such as source separation techniques, offer potential for improving specificity and accuracy in future studies, allowing for a more precise understanding of the underlying pathophysiology.[58,59].

Furthermore, the cross-sectional design of our study limits the ability to assess causal relationships or track the temporal progression of neuropathological changes in Parkinson’s disease. Additionally, the relatively small sample size may reduce the generalizability of our findings and decrease the statistical power to detect more subtle differences between groups. Another limitation is the use of diffusion-weighted MRI with low b-values, which may not fully capture the complexity of microstructural alterations in the brain. Future research incorporating longitudinal designs, larger cohorts, and more advanced diffusion MRI techniques—such as high angular resolution diffusion imaging (HARDI) or diffusion kurtosis imaging (DKI)—would enhance the ability to validate and expand upon our results. These approaches could provide a more detailed and comprehensive understanding of the neurobiological mechanisms underlying the heterogeneity of PD, helping to clarify the structural changes associated with different motor subtypes.

In conclusion, our multicontrast MRI study demonstrates distinct neuroimaging signatures of PD motor subtypes, with increased iron deposition in the substantia nigra of TD patients and more widespread white matter integrity changes in PIGD patients, especially in cerebellar pathways. These findings underscore the potential of neuroimaging biomarkers in elucidating the pathophysiological mechanisms underlying PD heterogeneity and may inform the development of targeted diagnostic and therapeutic strategies. By leveraging advanced MRI techniques and longitudinal designs, future research can further unravel the complex interplay between brain structure, function, and clinical phenotypes in PD, paving the way for a more personalized approach to managing this debilitating neurodegenerative disorder.

## Acknowledgments

The authors sincerely thank the patients and controls for their participation in the study. We also extend our gratitude to the staff at Hospital Santa Marta for their valuable assistance in collecting the MRI data.

## Author’s roles

1. Research Project: A. Conception, B. Organization, C. Execution; 2. Statistical Analysis: A. Design, B. Execution, C. Review, and Critique; 3. Manuscript Preparation: A. Writing the First Draft, B. Review and Critique.

DB: 1A-C, 2A-C, 3A; EN: 1B-C, 2A-B, 3A; PB: 1A-C, 2A-C, 3B; DP: 1C, 2A-C, 3B; TG: 1A-C, 2C, 3B; FM: 1C, 2C, 3B; NR: 1C, 2C, 3B; AF: 1C, 2C, 3B; XL: 1C, 2C, 3B; MT: 1A-C, 2C, 3B; FC: 1A-C, 2C, 3B.

## Data availability

A qualified investigator may obtain an anonymized dataset for the primary analysis provided in this paper upon reasonable request if the purpose is reproducing its results.

